# Rare variant burden complements polygenic risk in breast cancer and is associated with earlier diagnosis

**DOI:** 10.64898/2026.10.17.26365799

**Authors:** Zimeng Yu, Yikai Dong

## Abstract

Breast cancer affects approximately one in eight US women during their lifetime, and about 10% of new cases occur before age 45. Polygenic risk scores (PRS) are associated with breast cancer risk, and previous studies have evaluated PRS alongside pathogenic-variant carrier status. Whether a continuous, gene-effect-weighted rare-variant burden adds information beyond PRS, and whether the two genetic measures relate similarly to diagnosis age, remain unclear. We studied these questions using whole-genome sequencing and electronic health record data from roughly 8,000 breast cancer cases and 134,727 controls of genetically inferred European ancestry in the All of Us Research Program. In joint case-status models, increases in PRS and rare-variant burden (Burden_rare_) were associated with higher odds of breast cancer (odds ratios, 1.55 and 1.27, respectively). Adding both scores and their interaction to covariates increased the cross-validated area under the receiver operating characteristic curve from 0.745 to 0.782. The PRS odds ratio differed between noncarriers and carriers (1.56 versus 1.25), suggesting that their effects are not simply multiplicative. We then examined whether the same genetic measures were associated with recorded age at diagnosis among cases. In eligible cases, each standard deviation increase in Burden_rare_ was associated with diagnosis 1.82 years earlier, compared with 0.43 years earlier for PRS. The proportion carrying a qualifying rare variant decreased from 26.5% among cases diagnosed before age 35 to 3.2% among those diagnosed at age 70 or older. Higher breast cancer risk can itself produce an earlier mean diagnosis age among cases, so we compared observed age associations with those expected from age-scale risk models under age-constant hazard ratios. Notably, the PRS association was consistent with its risk-based expectation, whereas the Burden_rare_ association was stronger than expected. In this common whole-genome sequencing framework, the weighted rare-variant burden adds information beyond PRS for breast cancer case status and shows a more pronounced association with early diagnosis, guiding future studies of risk and diagnosis timing.

## Introduction

Both common and rare germline variation contribute to breast cancer susceptibility. Genome-wide association studies have identified many common risk loci, and a polygenic risk score (PRS) combines their effects. A score developed by the Breast Cancer Association Consortium from 313 variants was associated with breast cancer risk in prospective cohorts^[1]^. Rare pathogenic variation includes protein-truncating variants (PTVs) and missense variants classified as pathogenic or likely pathogenic in susceptibility genes. Large case-control studies have estimated elevated risks for variants in *BRCA1*, *BRCA2*, *PALB2*, *ATM*, *CHEK2*, and other genes, with the strength of association depending on the gene and variant class^[2,3]^.

Previous studies have estimated PRS associations within *BRCA1* and *BRCA2* carriers^[4,5]^, and carriers of other susceptibility-gene variants^[6]^. Population biobanks^[7,8]^ and larger case-control studies have also examined PRS and pathogenic-variant carrier status across broader samples^[9,10]^. These analyses generally classify rare variation by carrier status or gene. However, the added value of a continuous burden score that sums qualifying variants across genes beyond PRS for breast cancer case status remains uncertain. Joint modeling also allows us to test whether the PRS association with breast cancer case status differs between carriers and noncarriers.

Among women who develop breast cancer, age at diagnosis varies widely, and the genetic measures associated with case status may relate differently to earlier diagnosis. In US surveillance data, the median diagnosis age is 64 years, but about 10% of breast cancers are diagnosed before age 45^[11]^. Age-specific breast cancer risks have been estimated for pathogenic-variant carriers and for PRS^[3,8]^. A lower mean diagnosis age among cases with higher genetic scores does not by itself establish a stronger effect at younger ages, because an age-constant increase in breast cancer risk can produce the same pattern. It remains unclear whether PRS and a gene-weighted rare-variant burden have similar associations with age at diagnosis, or whether either association exceeds what its breast cancer risk association alone would predict under age-constant relative effects.

In this study, we used whole-genome sequencing and electronic health records from participants of genetically inferred European ancestry in the All of Us Research Program^[12,13]^ to evaluate the separate and joint associations of PRS and a continuous rare-variant burden weighted by external gene-specific effects (Burden_rare_) with breast cancer case status. Among cases, we examined the associations of both scores with recorded age at diagnosis. Both scores were independently associated with case status and improved breast cancer risk prediction; the PRS association was weaker in carriers than in noncarriers. Among cases, Burden_rare_ was more strongly associated with earlier diagnosis than PRS. Age-scale comparisons showed that the PRS age association was compatible with its risk-based expectation, whereas the burden association was stronger than expected. This joint analysis shows that the two sources of inherited risk contribute differently to case status and recorded diagnosis age.

## Methods

### Study population

We used the *All of Us* controlled-tier Curated Data Repository (CDR) version C2025Q4R6 and its short-read whole-genome sequencing (WGS) callsets^[13]^. Eligible participants were those recorded as female sex at birth with WGS data. *All of Us* applies sex-concordance checks before release, so we did not exclude participants again on inferred genetic sex. We restricted the analysis to genetically inferred European ancestry, using the program’s ancestry predictions and requiring a predicted European label with a maximum posterior probability above 0.75. The same rule was applied to cases and controls. We removed related individuals by keeping a maximal unrelated set among pairs with KING kinship >0.1^[14]^, and removed samples with genotype missingness above 3%. The flow of participants is shown in Figure 1.

**Figure 1.**
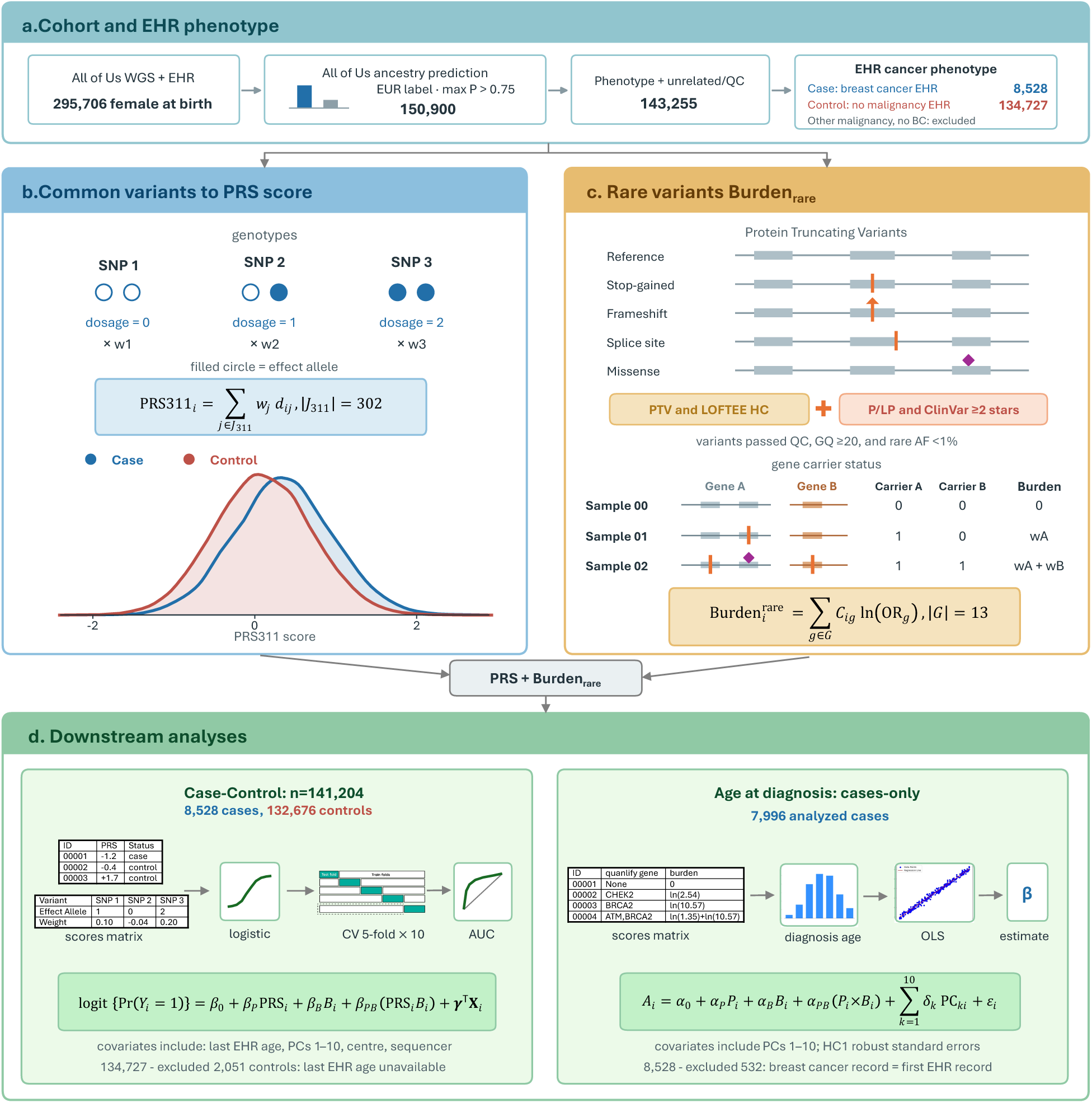
Cohort and genetic analysis workflow. (a) All of Us participants recorded female at birth with whole-genome sequencing and electronic health records underwent ancestry, relatedness, and quality control filtering, yielding 8,528 breast cancer cases and 134,727 controls. (b) PRS combines effect-allele dosages at 302 scored loci; curves show case and control distributions. (c) Schematic protein-truncating and pathogenic/likely pathogenic missense variants illustrate gene-level carrier indicators and the Burden_rare_. (d) Logistic regression assesses case status in 141,204 participants with repeated five-fold cross-validation; cases-only linear regression assesses age at diagnosis in 7,996 cases.

### Phenotypes

Cases were participants with at least one EHR condition record mapping to the SNOMED concept “Malignant tumor of breast” (OMOP concept 4112853) or any of its descendants. Controls were eligible participants without an EHR record of any malignant neoplasm (OMOP concept 443392 or its descendants); participants with another recorded malignancy but no qualifying breast cancer record were excluded. Recorded age at diagnosis was calculated from the earliest qualifying breast cancer record. Age at last EHR observation was the later of the last EHR record and the date of death. Case-status models required a non-missing age at last observation, excluding 2,051 participants, all controls. The age-at-diagnosis analysis additionally excluded 532 cases whose earliest breast cancer record was also their first EHR record, because the record date could reflect a diagnosis made before EHR follow-up began. In a sensitivity analysis, cases were required to have at least two qualifying records.

### Polygenic risk score

We calculated the PRS using PGS000004 from the PGS Catalog, which originally comprised 313 loci^[1,15]^, harmonised to GRCh38. Variants were matched to the *All of Us* array-and-common-allele-frequency (ACAF) callset by chromosome, position and alleles. Where strand or allele orientation was ambiguous, candidate records were resolved by comparing the cohort allele frequency with the published effect-allele frequency. In total, 304 of 313 variants were matched: 281 matched directly, 23 were resolved by allele frequency, one remained ambiguous and eight were absent. Missing genotypes were imputed using PLINK2 reference. Based on published evidence, we excluded two matched PRS loci in linkage disequilibrium with *CHEK2* c.1100delC^[16]^. This avoided counting that variant twice, since it also enters the rare-variant burden. Of the 311 loci remaining after this exclusion, 302 were matched and scored in All of Us. Scores were standardised within the analysis cohort. In sensitivity analyses we restricted the score to the 281 directly matched variants and repeated the scoring without mean imputation. For participant i, the score was the weighted sum of effect-allele dosages across retained loci:

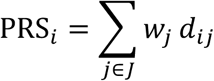

Here J is the set of matched loci after removing the two CHEK2-tagging variants, wⱼ is the per-effect-allele log odds ratio for breast cancer estimated in the original PRS development data and supplied with PGS000004^[1,15]^, rather than re-estimated in All of Us, and dᵢⱼ is the effect-allele dosage (0, 1 or 2), replaced by the cohort mean dosage when missing.

### Rare-variant burden

The burden panel comprised 13 breast cancer susceptibility genes for which Rowlands et al. reported combined pathogenic-variant odds ratios: *ATM*, *BARD1*, *BRCA1*, *BRCA2*, *CHEK2*, *PALB2*, *RAD51C*, *RAD51D*, *TP53*, CDH1, NF1, PTEN and *STK1*1^[17]^. Qualifying PTVs were stop-gained, frameshift, splice-donor and splice-acceptor variants with both the qualifying consequence and high-confidence LOFTEE annotation on the same retained transcript of the same gene^[18]^; eligibility was not restricted to the canonical transcript. Qualifying missense variants required germline pathogenic/likely pathogenic classification in ClinVar (13 September 2026 release), at least two review stars, and relevant disease and inheritance assertions. Low-penetrance variants, risk alleles, conflicting classifications and opposing benign/likely benign assertions were excluded; unresolved records were held out. Both classes required a known, minor allele frequency below 1% in the *All of Us* European-ancestry callset. We required variants to pass site- and genotype-level quality filters. Carrier genotypes required genotype quality ≥ 20 and target-alternate allele fraction 0.30–0.80 for heterozygous or ≥ 0.80 for homozygous-alternate calls. Allele fraction was the target-alternate allelic depth divided by total allelic depth after normalized-allele matching. Missing required QC failed inclusion; no blanket read-depth threshold was applied. A carrier had at least one qualifying PTV or missense variant in any panel gene.

Burden_rare_ was the sum, over panel genes in which a participant carried a qualifying variant, of the gene’s log odds ratio. Gene weights came from the combined PTV and non-PTV pathogenic-variant estimates in Rowlands et al.^[17]^. These are approximate external gene-level weights because the source variant definitions differ from ours. Multiple qualifying variants within one gene contributed once; variants in different genes contributed separately. To assess the imprecise STK11 weight (OR 1.60, 95% CI 0.48–5.30), we compared the full panel with a twelve-gene panel excluding STK11, using the *same* participants, PRS, covariates and cross-validation folds. Explicitly, the participant-level score was:

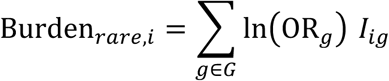

G is the set of 13 panel genes, ORg is the published combined pathogenic-variant odds ratio for gene g, and Iᵢg is an indicator that equals 1 when participant i carries at least one qualifying PTV or missense variant in that gene and 0 otherwise. Multiple qualifying variants within a gene therefore contributed only once.

### Statistical analysis

#### Breast cancer risk models

We fitted logistic regression models of case status on standardised PRS, standardised Burden_rare_ and their product. Covariates were age at last EHR observation, the first ten genetic principal components (PCs), sequencing centre and sequencer model. Sequencing centre identifies the whole-genome sequencing site, and sequencer model identifies the instrument used; we adjusted for both to account for potential technical differences in genotype measurement. Five models were compared: covariates only, covariates plus PRS, covariates plus Burden_rare_, an additive model with both scores, and an interaction model with both scores and their product. Coefficients are reported from fits to the full cohort, with heteroskedasticity-robust (HC1) standard errors. Breast cancer risk prediction was evaluated with 5-fold cross-validation repeated 10 times, stratified jointly on case status and carrier status. For each repeat we computed the out-of-fold AUC of every model and compared paired models with DeLong’s test^[19]^. We report mean AUC and ΔAUC over repeats, with DeLong confidence intervals and P values from the first repeat; these intervals are not confidence intervals for the repeated-CV means. Full-cohort fits use sample standard deviations; cross-validation standardization uses training-fold parameters. The logistic model has no tuning parameters and we did no model selection, so nested cross-validation was not needed. To describe the joint distribution, we compared observed case proportions by PRS decile and carrier status for the full panel with expectations from a logistic model without interaction, adjusted for continuous PRS, carrier status, age at last EHR observation and PCs 1–10. This descriptive comparison omitted sequencing centre and sequencer model; the reported genetic odds ratios came from the models adjusted for those technical factors.

The full interaction model was:

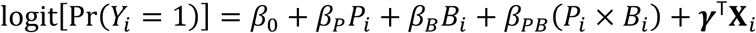

Here Yᵢ denotes case status; Pᵢ and Bᵢ are the standardised PRS and Burden_rare_; and Xᵢ contains age at last EHR observation, PCs 1–10, sequencing centre and sequencer model. Four nested models were fitted by setting selected coefficients to zero: β_P_=β_B_=β_PB_=0 for covariates only; β_B_=β_PB_=0 for covariates plus PRS; β_P_=β_PB_=0 for covariates plus Burden_rare_; and β_PB_=0 for the additive PRS and Burden_rare_ model. The β_PB_ coefficient measures departure from multiplicative odds in the full model.

#### Age at diagnosis among cases

We fitted linear models of recorded age at diagnosis, in years, among eligible cases. All models included PCs 1–10; the four additional specifications included PRS alone, Burden_rare_ alone, both scores additively, or both scores and their interaction. We used HC1 robust standard errors and measured each score’s contribution as the reduction in R² when that score was removed from the additive model. We then compared the observed age coefficients with those expected from age-scale Cox models under age-constant hazard ratios, since a score associated with higher risk can also be associated with earlier diagnosis among cases without a stronger effect at younger ages (Supplementary Methods). The full interaction model was:

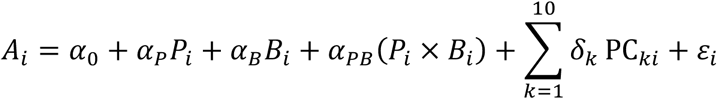

Here Aᵢ is recorded age at diagnosis for case i; Pᵢ and Bᵢ are standardised within the age-at-diagnosis analysis cohort; and PCₖᵢ is principal component k for case i. The PCs-only, PRS-only, Burden_rare_-only and additive models omit the corresponding score and interaction terms from this equation. In the additive model, α_P_ and α_B_ are adjusted differences in recorded diagnosis age, in years, per standard deviation of each score. The α_PB_ term measures departure from an additive age association in the full model.

#### Case-control interaction

We examined whether the PRS association with breast cancer case status differed by rare-variant carrier status. Participants were divided into non-carriers and carriers of qualifying variants in the burden panel. Within each stratum s, we fitted a logistic model with PRS standardised in the full case-control cohort:

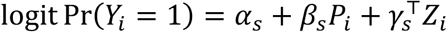

Here Yᵢ=1 denotes breast cancer case status, Pᵢ is the standardised PRS, and Zᵢ contains age at last EHR observation, PCs 1–10, sequencing centre and sequencer model. The stratum-specific coefficient βₛ is the log odds ratio per standard deviation of PRS. We used HC1 robust standard errors and report *exp(β̂_s_*)with 95% confidence intervals. For carriers (c) versus non-carriers (n), we calculated the ratio of PRS odds ratios and the standard error of the difference in log odds slopes as follows:

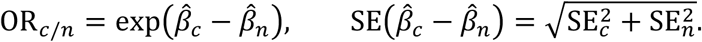

We also fitted age-scale Cox models within the same carrier strata to estimate the PRS association with the hazard of a first breast cancer record:

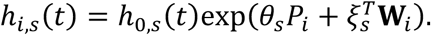

Here t is attained age, h0,s(t) is the stratum-specific baseline hazard, and Wᵢ contains PCs 1–10, sequencing centre and sequencer model; exp(θₛ) is the hazard ratio per standard deviation of PRS. Follow-up began at age 18 or one year after the first EHR record, whichever was later, and ended at the first breast cancer record, last EHR observation or death. Figure 6a presents the stratum-specific logistic odds ratios and Cox hazard ratios with 95% confidence intervals.

#### Age-scale models and expected age-at-diagnosis associations

We first estimated the case-only age coefficient expected from a genetic association with breast cancer risk that remains constant on the hazard-ratio scale across age. We fitted a Cox model with attained age t as the time scale and delayed entry. Follow-up began at age 18 or one year after the first EHR record, whichever was later, and ended at the first breast cancer record, last EHR observation or death:

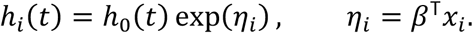

In this model, xᵢ is the covariate vector, β contains its log hazard coefficients, and ηᵢ is the fitted linear predictor. For a case with a first breast cancer record at age tᵢ, R(tᵢ) is the set of participants still under observation and at risk at that age. We calculated the hazard-weighted mean covariate vector m(tᵢ) in that risk set and expressed the case’s observed covariates as m(tᵢ) plus a residual rᵢ:

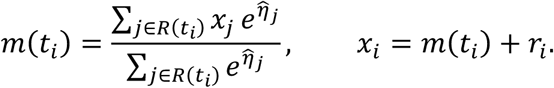

Let Z contain the centred score and PC predictors for the additive case-only age regression in Table 3, excluding the intercept, and let t̄ be mean recorded age at diagnosis among cases. The comparison concerns the regression slopes rather than the intercept. Replacing each case’s predictors by their hazard-weighted risk-set means gives the slopes expected under the fitted age-constant-hazard model. The observed slope vector can be written as that expected component plus a remainder:

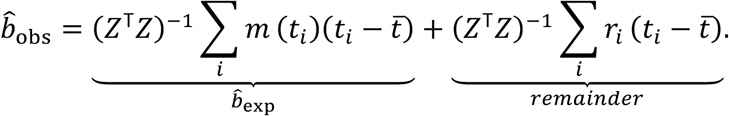

The first term is the expected case-only coefficient. The remainder reflects how case covariates differ from their hazard-weighted risk-set means across diagnosis ages and is related to the Schoenfeld-residual score for an age-varying effect. We compared observed and expected coefficients before examining age variation directly with an age-scale Cox model whose genetic coefficient could differ across attained-age bands:

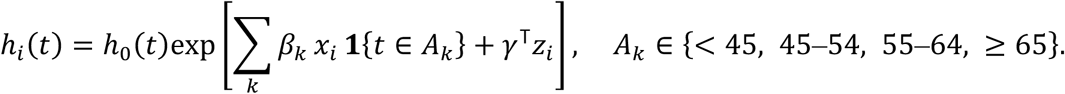

Here Aₖ denotes the attained-age bands <45, 45–54, 55–64 and ≥65 years; βₖ is the log hazard coefficient for genetic measure xᵢ in band k, and zᵢ contains adjustment covariates. We estimated band-specific hazard ratios to assess how the genetic association varied with age. We then examined whether depletion of higher-risk women from later-age risk sets could account for a declining population-level hazard ratio even if the individual-level effect remained constant. For this analysis, we multiplied each individual hazard by a gamma frailty uᵢ:

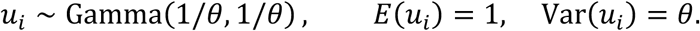

Gamma frailty was parameterised by shape and rate, giving mean 1 and variance θ. We bounded θ using the age pattern of PRS among non-carriers, then compared frailty-adjusted expected age-at-diagnosis coefficients with the observed coefficients (Supplementary Methods).

#### Sensitivity analyses

We repeated the case-status and age-at-diagnosis analyses with 12-gene panels that excluded STK11 or CDH1, separately. We also restricted qualifying variants to PTVs, tightened the upper allele-fraction threshold for heterozygous TP53 and NF1 calls to 0.70, and lowered the ClinVar review threshold to at least one star. We compared these specifications with the historical nine-gene PTV score and staged updates to its weights, genotype quality control and eligible missense variants; the historical scores retained their original definitions. For case-status prediction, we recalculated out-of-fold ROC curves and AUCs using the repeated cross-validation procedure described above (Supplementary Figures). We also repeated the comparison of earlier and later diagnosis among cases using <40 versus ≥40 years as the threshold.

#### Disclosure control

In line with the *All of Us* Data and Statistics Dissemination Policy, no count between 1 and 20 participants (or between 1 and 39 alleles) is reported, either directly or in a form from which it could be derived. Affected cells are shown as ≤ 20 and, where needed, a second cell is suppressed so that the first cannot be recovered by subtraction. Test statistics whose inputs include such a cell are not reported.

#### Software

Analyses used Python with pandas, statsmodels and scikit-learn; Hail 0.2.137; PLINK 2^[20]^; and bcftools. Analysis code is available at https://github.com/martina-yu/AoU_Comr.

## Results

### Study cohort

Among 295,706 women with WGS, 150,900 had genetically inferred European ancestry. Relatedness and sample QC left 143,255 participants: 8,528 cases and 134,727 controls (Figure 1). Mean recorded age at diagnosis among cases was 59.9 years (SD 11.8). Cases were older at enrolment than controls (64.0 versus 51.2 years; Table 1). Of 141,204 participants in the case-status model, 3,209 (2.3%) carried a qualifying PTV or missense variant in a panel gene (Figure 2).

**Figure 2.**
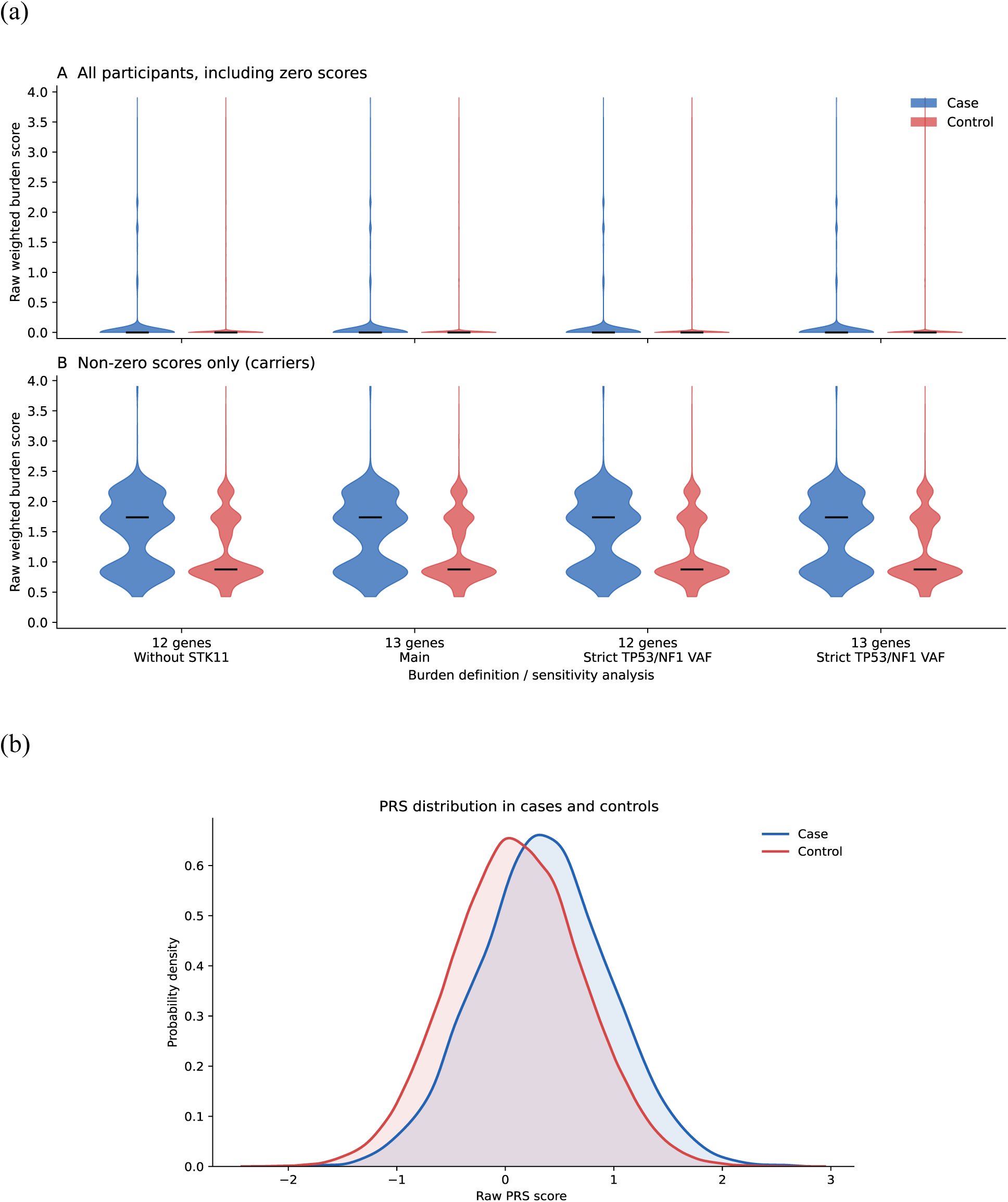
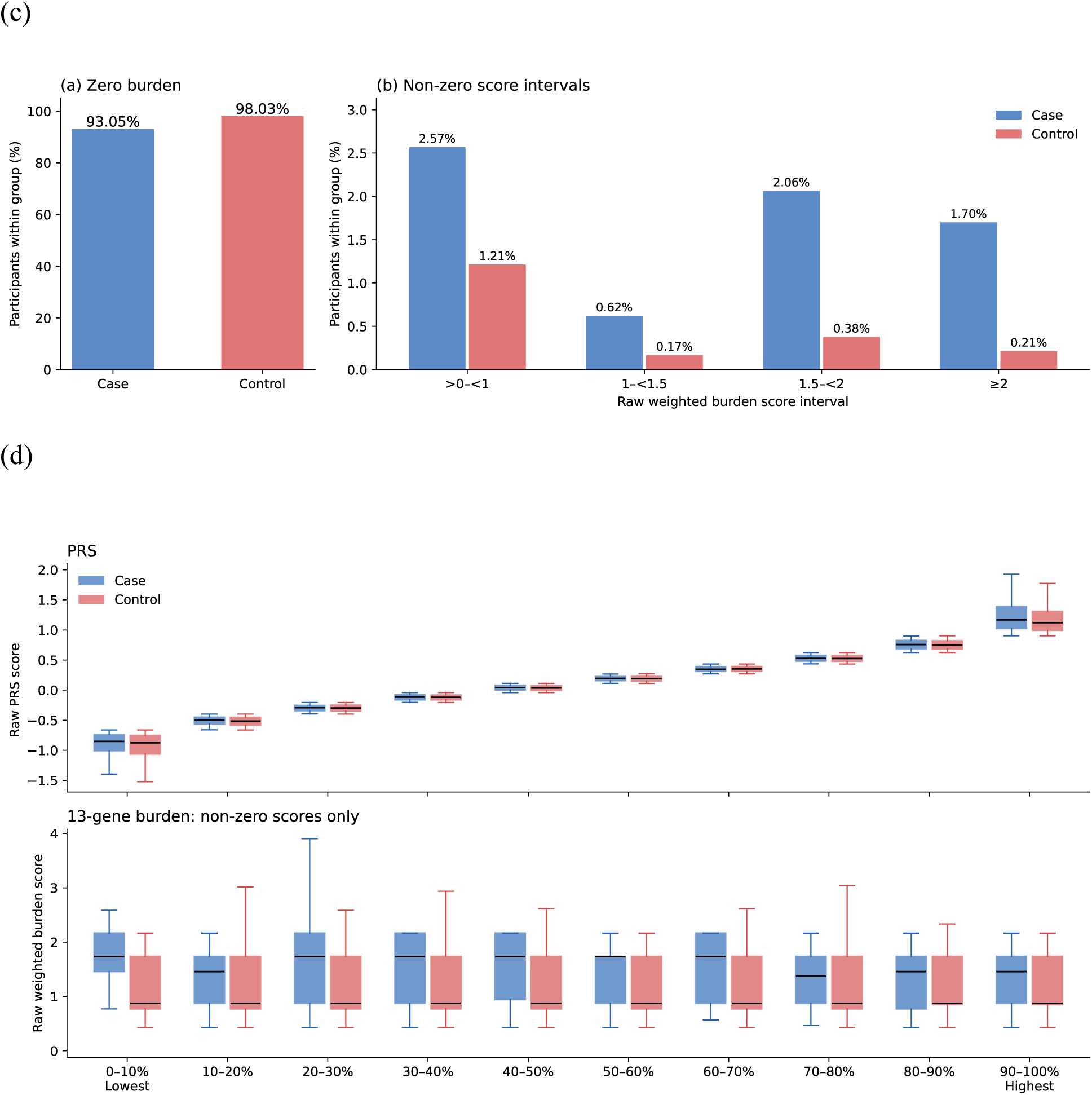
Genetic score distributions in the case-control cohort (n=141,204). (a) Burden_rare_ distributions under the definitions shown, for all participants (top) and carriers (bottom); black bars mark medians. (b) Raw PRS density by case status. (c) Percentages with zero and positive Burden_rare_ within cases and controls. (d) Raw PRS and nonzero Burden_rare_ across full-cohort PRS deciles. Cases are blue and controls red.

**Table 1.** Characteristics of the study cohort.

|  | Case-status cases | Case-status controls | Age-analysis cases |
| --- | --- | --- | --- |
| N | 8,528 | 134,727 | 7,996 |
| Age at diagnosis, years, mean (SD) | 59.9 (11.8) |  | 60.2 (11.8) |
| Age at diagnosis, years, median [IQR] | 60.7 [51.5-68.4] |  | 61.1 [51.9-68.7] |
| Age at enrollment, years, mean (SD) | 64.0 (11.6) | 51.2 (16.8) | 64.1 (11.6) |
| Age at last EHR observation, years, mean (SD) | 68.1 (11.7) | 53.9 (17.0) | 68.2 (11.7) |
| BMI, kg/m<sup>2</sup>, mean (SD) | 28.5 (6.7) | 29.4 (7.9) | 28.5 (6.8) |
| PRS (raw score), mean (SD) | 0.4 (0.6) | 0.1 (0.6) | 0.4 (0.6) |
| BMI missing, n (%) | 92 (1.1) | 5,107 (3.8) | suppressed (secondary) |
| Age at last EHR observation missing, n (%) | 0 (0.0) | 2,051 (1.5) | 0 (0.0) |
| Menopausal status, n (%) |  |  |  |
| Premenopausal | 620 (7.3) | 49,735 (36.9) | 592 (7.4) |
| Postmenopausal | 7,610 (89.2) | 75,983 (56.4) | 7,133 (89.2) |
| Uncertain or unknown | 149 (1.7) | 4,801 (3.6) | suppressed (secondary) |
| Missing | 149 (1.7) | 4,208 (3.1) | suppressed (secondary) |
| Smoking status, n (%) |  |  |  |
| Never | 4,932 (57.8) | 80,136 (59.5) | 4,612 (57.7) |
| Former or unknown currency | 2,974 (34.9) | 35,699 (26.5) | 2,796 (35.0) |
| Current | 442 (5.2) | 13,614 (10.1) | suppressed (secondary) |
| Missing | 180 (2.1) | 5,278 (3.9) | suppressed (secondary) |
| Self-reported race, n (%) |  |  |  |
| White | 8,117 (95.2) | 126,790 (94.1) | 7,611 (95.2) |
| Other, multiple, or not reported | 411 (4.8) | 7,937 (5.9) | 385 (4.8) |
| Sequencing center, n (%) |  |  |  |
| Broad Institute | 4,963 (58.2) | 79,319 (58.9) | 4,639 (58.0) |
| University of Washington | 997 (11.7) | 16,069 (11.9) | 936 (11.7) |
| Baylor College of Medicine | 2,568 (30.1) | 39,339 (29.2) | 2,421 (30.3) |
| Sequencer, n (%) |  |  |  |
| NovaSeq6000 S4 | 8,293 (97.2) | 127,580 (94.7) | suppressed (secondary) |
| NovaSeq X 25B | 235 (2.8) | 7,147 (5.3) | suppressed (secondary) |

Counts between 1 and 20, and cells from which such a count could be derived, are suppressed.

### Breast cancer case status

In the joint logistic model, the odds ratio for breast cancer case status per standard deviation was 1.55 (95% CI 1.52–1.59) for PRS and 1.27 (1.25–1.29) for Burden_rare_, each evaluated at the other score’s mean. Their interaction was negative on the multiplicative odds scale (OR 0.972, 95% CI 0.958–0.985; P=6.0×10⁻⁵; Table 2).

**Table 2.** Joint logistic model of case status and cross-validated breast cancer risk prediction. Models adjust for age at last EHR observation, PC1–10, sequencing centre and sequencer. AUC and ΔAUC are means over ten repeats; 95% CIs and paired DeLong P values are from the first repeat and are not confidence intervals for the repeated-CV means. n=141,204 (8,528 cases).

|  | Estimate (95% CI) | P |
| --- | --- | --- |
| **A. Joint model, full cohort (OR per SD)** |  |  |
| PRS | 1.55 (1.52–1.59) | $< 10^{-300}$ |
| Burden<sub>rare</sub> | 1.27 (1.25–1.29) | $6.9 \times 10^{-233}$ |
| PRS × Burden<sub>rare</sub> | 0.972 (0.958–0.985) | $6.0 \times 10^{-5}$ |
| **B. Out-of-fold AUC, 10 × 5-fold CV (mean; 95% CI from repeat 1)** |  |  |
| Covariates-only | 0.7452 (0.7406–0.7498) | – |
| PRS-only | 0.7682 (0.7637–0.7728) | – |
| Burden<sub>rare</sub>-only | 0.7588 (0.7543–0.7633) | – |
| PRS+Burden<sub>rare</sub> (no interaction) | 0.7813 (0.7770–0.7858) | – |
| PRS+Burden<sub>rare</sub> + Interaction (Joint) | 0.7815 (0.7771–0.7859) | – |
| **C. ΔAUC (mean over repeats; repeat-1 DeLong 95% CI and P)** |  |  |
| Interaction – Additive | 0.000181 (0.000023–0.000314) | $2.303 \times 10^{-2}$ |
| Additive – Cov | 0.036150 (0.033258–0.039109) | $8.28 \times 10^{-130}$ |
| Additive – PRS | 0.013098 (0.011202–0.015021) | $2.81 \times 10^{-41}$ |
| Additive – Burden | 0.022523 (0.020250–0.024841) | $1.40 \times 10^{-82}$ |
| Interaction – Cov | 0.036331 (0.033380–0.039323) | $4.75 \times 10^{-127}$ |
| Interaction – PRS | 0.013279 (0.011326–0.015233) | $1.68 \times 10^{-40}$ |
| Interaction – Burden | 0.022704 (0.020388–0.025039) | $1.13 \times 10^{-81}$ |
| PRS – Cov | 0.023052 (0.020712–0.025432) | $7.58 \times 10^{-82}$ |
| Burden – Cov | 0.013627 (0.011661–0.015615) | $1.18 \times 10^{-41}$ |

Cov: covariates only; PRS: covariates + PRS; Burden: covariates + Burden; Additive: covariates + Burden + PRS; Interaction: Additive + Burden × PRS

**Table 3.** Associations of Burden*_rare_* and PRS with recorded age at breast cancer diagnosis among cases (n = 7,996).

| **A. Joint OLS model** |  |  |
| --- | --- | --- |
| Genetic measure | Estimate, years per SD (95% CI) | P value |
| $\text{Burden}_{\text{rare}}$ | −1.82 (−2.11 to −1.53) | $1.5 \times 10^{-35}$ |
| PRS | −0.43 (−0.68 to −0.18) | 0.00075 |
| **B. Explained variance** |  |  |
| Model | R<sup>2</sup> | — |
| PCs only | 0.20% | — |
| PRS + PCs | 0.26% | — |
| $\text{Burden}_{\text{rare}}$ + PCs | 2.49% | — |
| Joint: PRS + $\text{Burden}_{\text{rare}}$ + PCs | 2.63% | — |
| **C. Incremental explained variance** |  |  |
| Score added | Reduced model | $\Delta R^2$, percentage points |
| $\text{Burden}_{\text{rare}}$ | PRS + PCs | 2.37 |
| PRS | $\text{Burden}_{\text{rare}}$ + PCs | 0.13 |

The covariate-only model had a mean out-of-fold AUC of 0.745, driven largely by age at last EHR observation. Adding PRS increased AUC by 0.023, and adding Burden_rare_ increased it by 0.014 (Figure 3). The model with both scores and their interaction reached 0.782, an increase of 0.036 over covariates alone and 0.01328 over covariates plus PRS. In the first cross-validation repeat, the latter increase was 0.01328 (95% CI 0.01133–0.01523; paired DeLong P=1.7×10⁻⁴⁰); AUCs varied by less than 0.0003 across ten repeats. Because age at last observation reflects both attained age and the length of EHR follow-up, we interpret the change in AUC rather than the absolute value as the contribution of the genetic scores.

**Figure 3.**
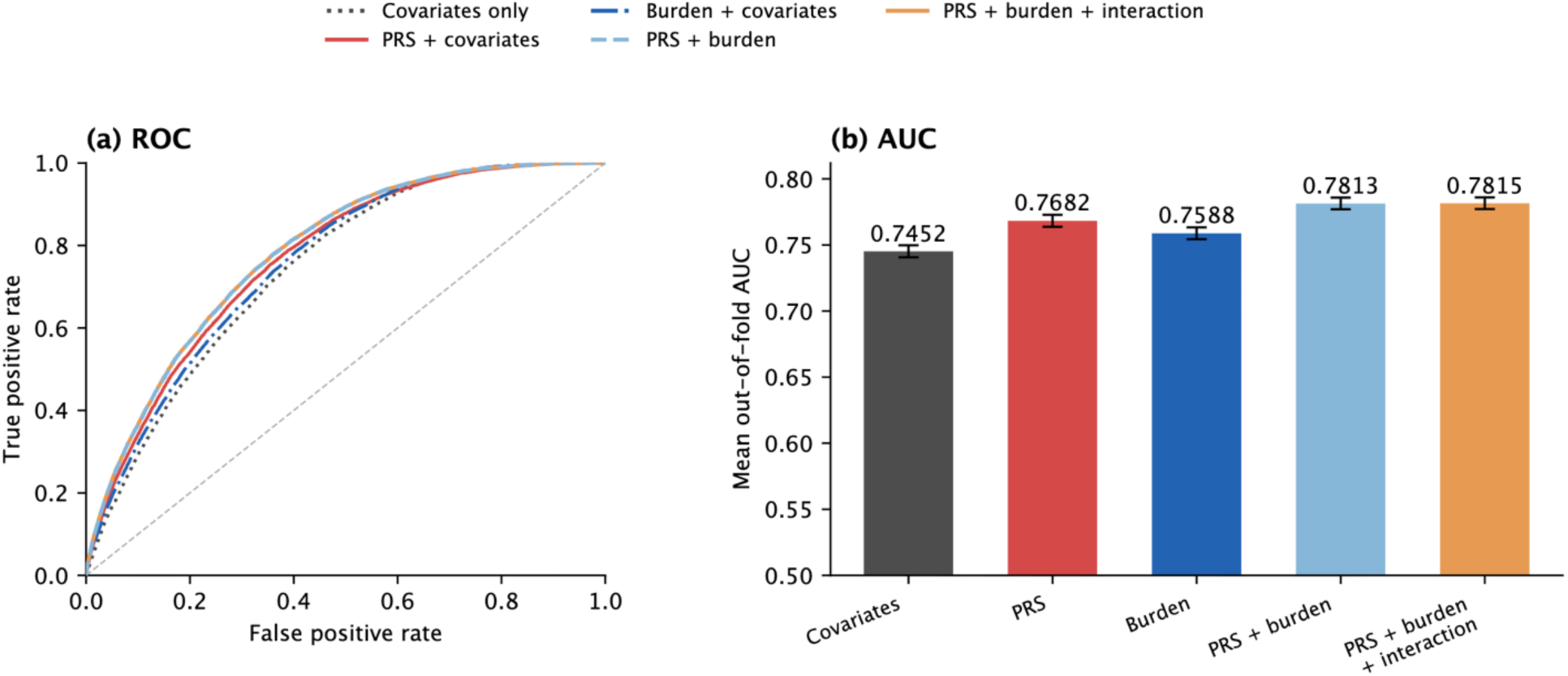
Breast cancer risk model performance using PRS and Burden_rare_. (a) ROC curves from out-of-fold predictions in the first five-fold cross-validation repeat for covariates alone, covariates plus PRS, covariates plus Burden_rare_, covariates plus both scores, and covariates plus both scores and their interaction. (b) Mean out-of-fold AUC across ten repeats of five-fold cross-validation; error bars show DeLong 95% confidence intervals from the first repeat.

### Joint distribution of PRS and rare-variant carrier status

The observed case proportion rose across PRS deciles among noncarriers and was higher among carriers at every decile. Compared with the model without interaction, carrier cases were less frequent than expected in the upper PRS deciles and more frequent than expected in the lower deciles. This descriptive pattern accords with the negative interaction estimate in Table 2.

### Recorded age at diagnosis among cases

Figure 4 shows the distribution of recorded diagnosis age across PRS groups and carrier status among 7,996 cases. PRS quartiles, defined in the full case-control cohort, were represented across all age bands (Figure 4a). The carrier proportion fell from 26.5% (95% CI 20.1–33.9%) among cases diagnosed before age 35 to 3.2% (2.5–4.2%) among those diagnosed at age 70 or older (Figure 4b). In both the median-split and extreme-PRS comparisons, carriers tended to have younger recorded diagnosis ages than noncarriers (Figure 4c).

**Figure 4.**
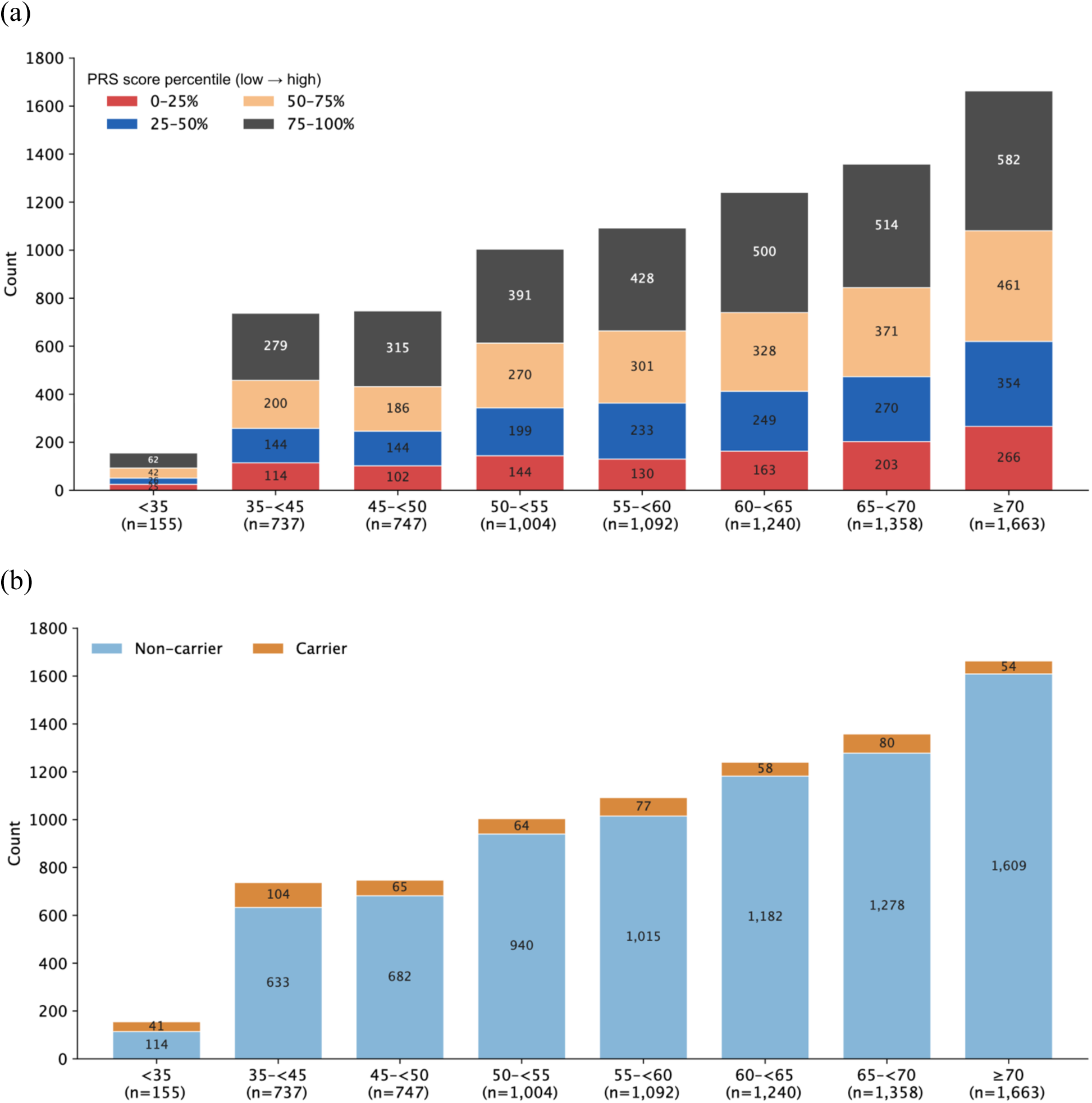
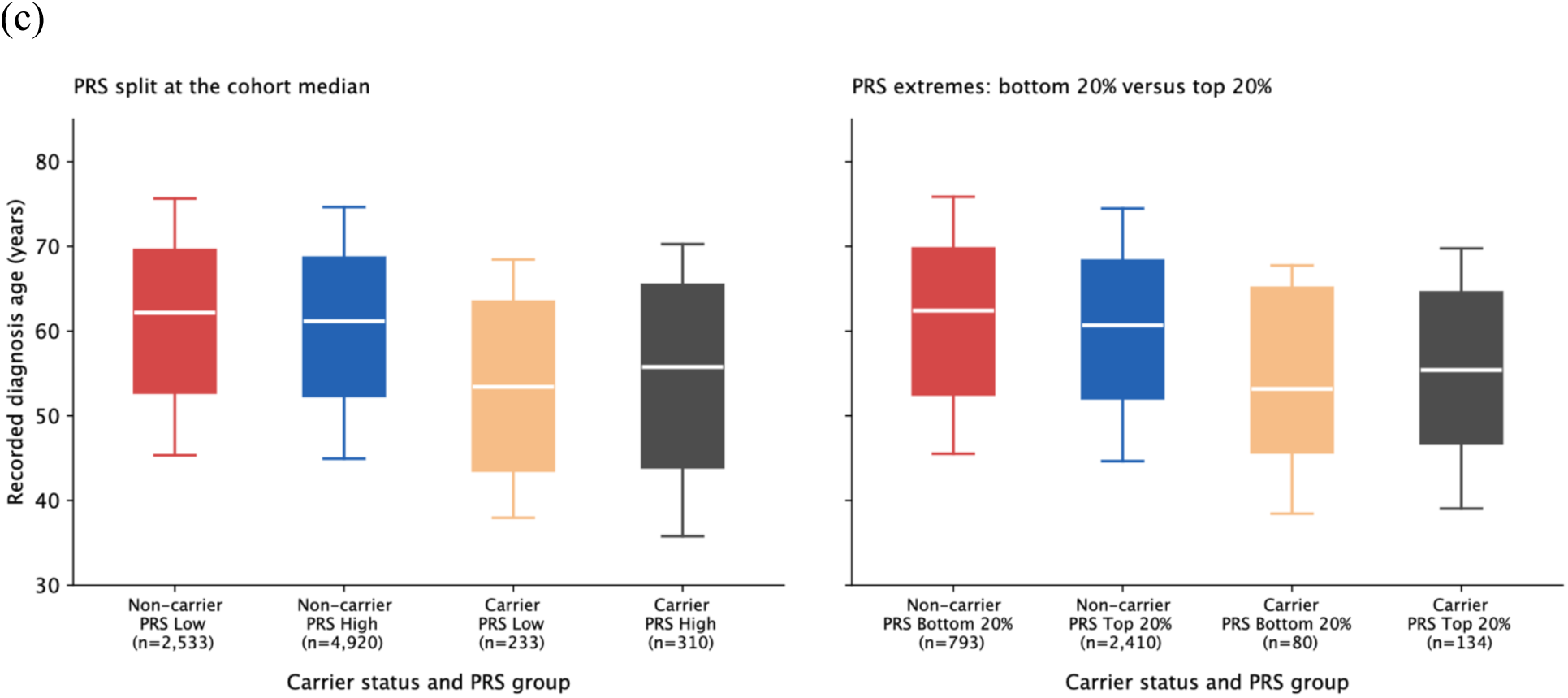
Recorded age at diagnosis by PRS and rare-variant carrier status among cases (n=7,996). (a) Counts of cases in each diagnosis-age band by PRS quartile; quartile cut points were defined in the full case-control cohort. (b) Counts of carriers and non-carriers in the same age bands. (c) Box plots of recorded age at diagnosis by carrier status and PRS group: the left panel splits PRS at the full-cohort median, and the right panel compares the bottom and top 20% of the full-cohort PRS distribution.

In the additive case-only model, each standard deviation increase in Burden_rare_ was associated with diagnosis 1.82 years earlier (95% CI 1.53–2.11 years earlier; P=1.5×10⁻³⁵), compared with 0.43 years earlier for PRS (0.18–0.68 years earlier; P=0.00075; Table 3). Together, the scores explained 2.63% of variation in recorded diagnosis age; removing Burden_rare_ reduced R² by 2.37 percentage points. Figure 5 compares the case-status and diagnosis-age estimates across rare-variant definitions.

**Figure 5.**
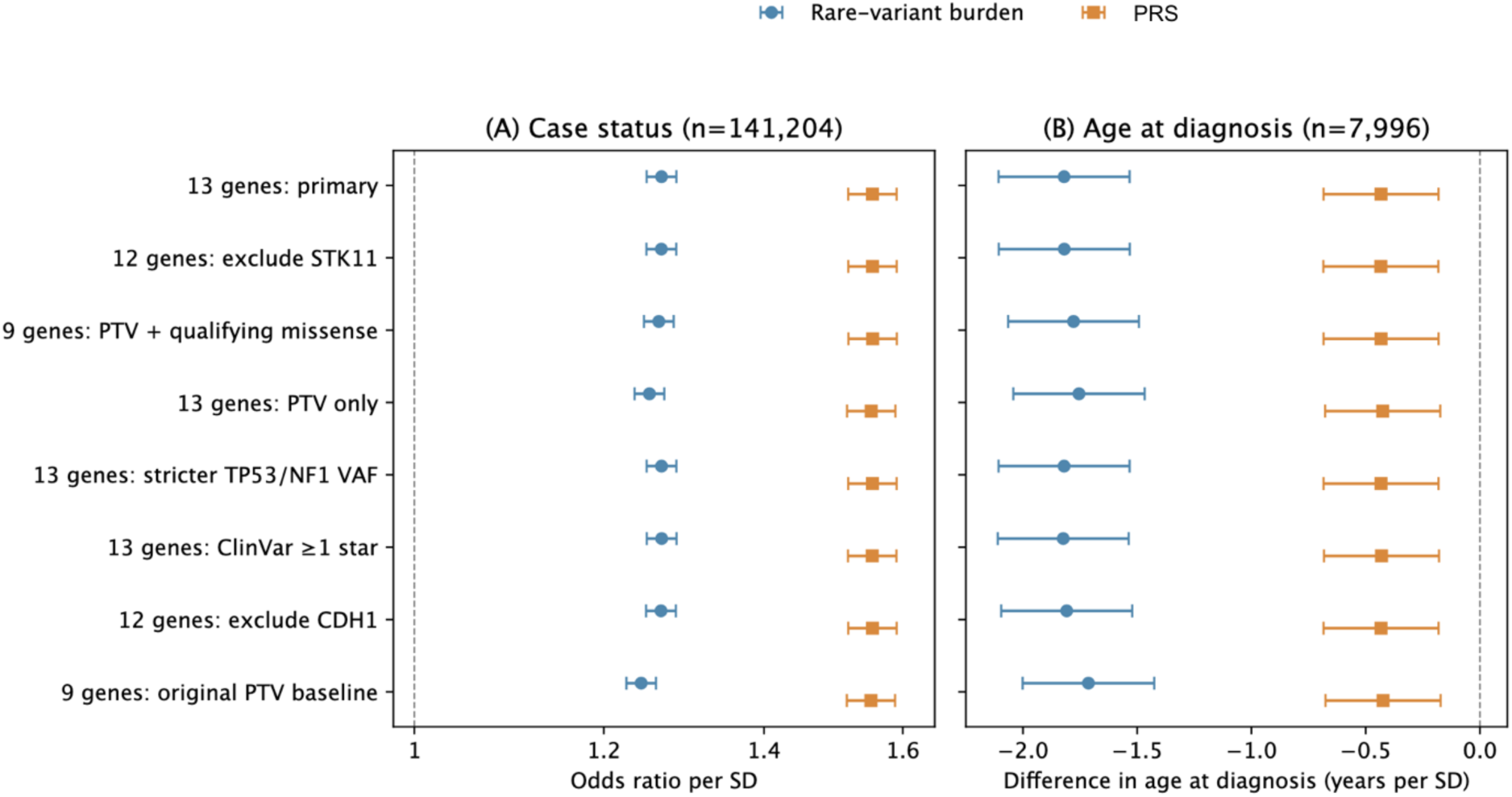
Case-status and diagnosis-age associations across rare-variant definitions. (a) Case-status odds ratios per standard deviation from the joint logistic model, including PRS, Burden_rare_, their interaction, age at last EHR observation, PC1–10, sequencing centre and sequencer model (n=141,204). (b) Difference in recorded age at diagnosis per standard deviation among cases from an OLS model including PRS, Burden_rare_ and PC1–10, without interaction (n=7,996). Points show Burden_rare_ in blue circles and PRS in orange squares; horizontal lines show 95% confidence intervals based on HC1 robust standard errors.

**Figure 6.**
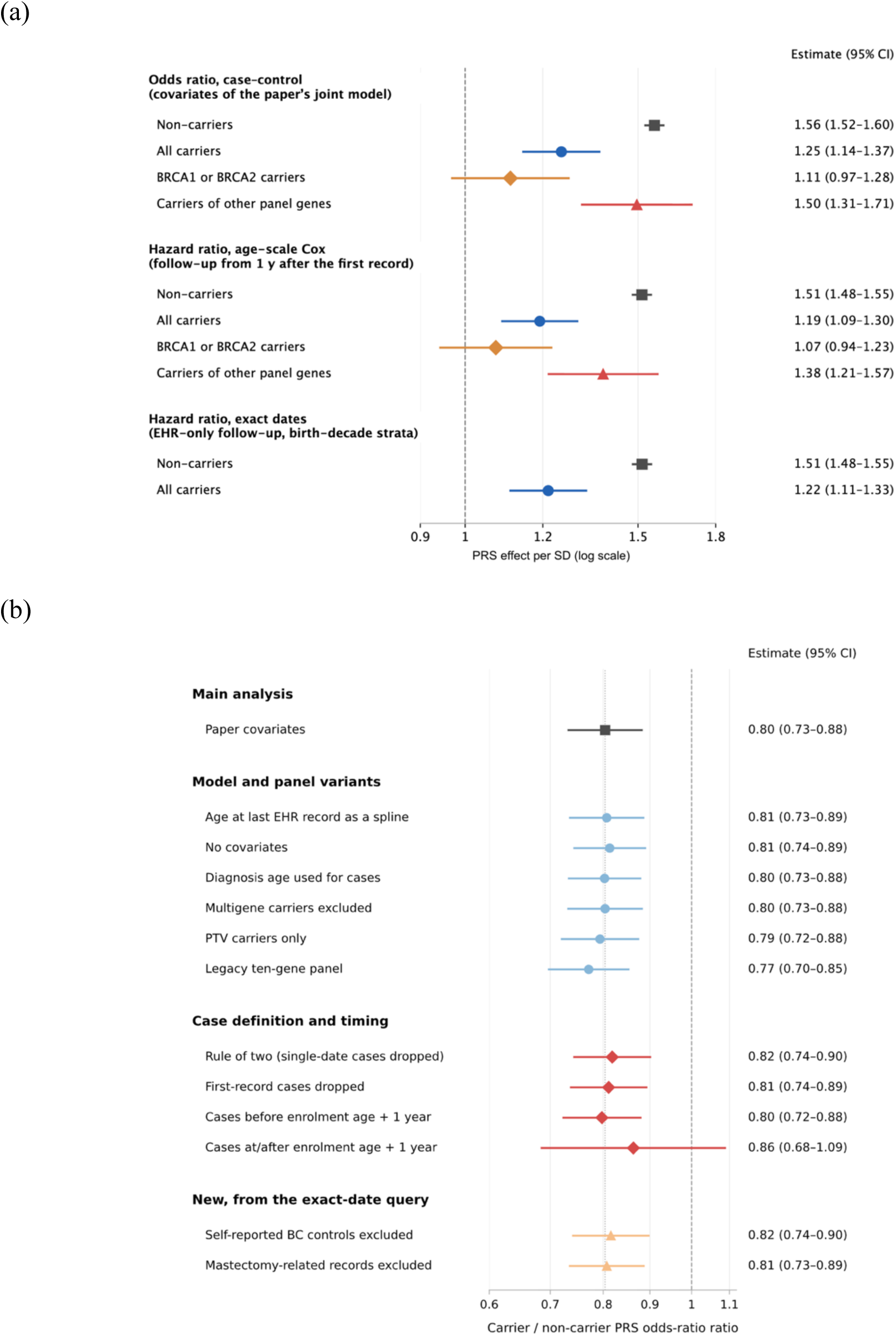
PRS associations with breast cancer risk by carrier status. (a) Odds ratios (ORs) per standard deviation of PRS from logistic models of breast cancer case status fitted separately in noncarriers and carriers, with estimates for BRCA1/BRCA2 and other panel-gene carriers. Age-scale Cox hazard ratios (HRs) are shown below the logistic estimates; an exact-date EHR follow-up sensitivity analysis appears at the bottom. (b) Ratios of the logistic PRS OR in carriers to that in noncarriers across the model, gene-panel and case-definition specifications shown. Points and horizontal lines indicate estimates and 95% confidence intervals; the vertical line in (b) marks a ratio of 1.

Case-status odds ratios and case-only age coefficients changed little across the panel, variant-filter and score-definition analyses (Figure 5a,b). Excluding STK11 changed the joint-model AUC from 0.78151 to 0.78149, the Burden_rare_ odds ratio from 1.269 to 1.268, and its age coefficient from −1.820 to −1.819 years per standard deviation. Across the alternative burden specifications, PRS odds ratios remained near 1.55, Burden_rare_ odds ratios ranged from 1.25 to 1.27, and age coefficients ranged from −0.44 to −0.43 years for PRS and from −1.82 to −1.75 years for Burden_rare_. The historical nine-gene PTV score is shown separately in Figure 5 because its weights and eligibility rules differed. Supplementary Tables S3 and S5 report the AUC estimates and paired increments.

We then assessed PRS associations within carrier strata. In separate logistic models, the PRS odds ratio was 1.56 (95% CI 1.52–1.60) in noncarriers and 1.25 (1.14–1.37) in carriers (Figure 6a). The corresponding age-scale Cox hazard ratios were 1.51 (1.48–1.55) and 1.19 (1.09–1.30); an exact-date EHR follow-up analysis gave similar estimates. The carrier-to-noncarrier ratio of logistic PRS odds ratios was 0.80 (0.73–0.88), with the same direction across model, panel and case-definition checks (Figure 6b). The exploratory PRS-by-carrier association with recorded diagnosis age was imprecise (interaction +0.98 years per PRS SD, 95% CI −0.08 to 2.04). In case-status models, PRS interactions differed across seven gene categories (heterogeneity P=0.0126); the BRCA1 interaction was the most negative (OR 0.652, 95% CI 0.514–0.828; q=0.0030), while the BRCA2 estimate was 0.786 (0.640–0.965; q=0.0758; Supplementary Table S7 and Figure S1).

### Age-scale expectations and age-specific hazard ratios

We next compared the observed case-only age coefficients with those expected if the genetic hazard ratios remained constant across age (Figure 7a). For PRS, the expected association was 0.23–0.30 years earlier per standard deviation, compatible with the observed 0.43 years earlier (P=0.29). For Burden_rare_, the expected association was 0.72–0.80 years earlier, substantially smaller than the observed 1.82 years earlier (P=3×10⁻¹⁴). In age-band Cox models, the PRS hazard ratio changed little with attained age (ratio per decade, 1.00), whereas the BRCA1/BRCA2 carrier hazard ratio declined from 8.4 before age 45 to 2.7 at age 65 or older (Figure 7b). Gamma-frailty simulations suggested that depletion of higher-risk women from later risk sets could account for much of the carrier hazard-ratio decline, but the observed Burden_rare_ age coefficient remained beyond the frailty-adjusted expectation (Figure 7a,b).

**Figure 7.**
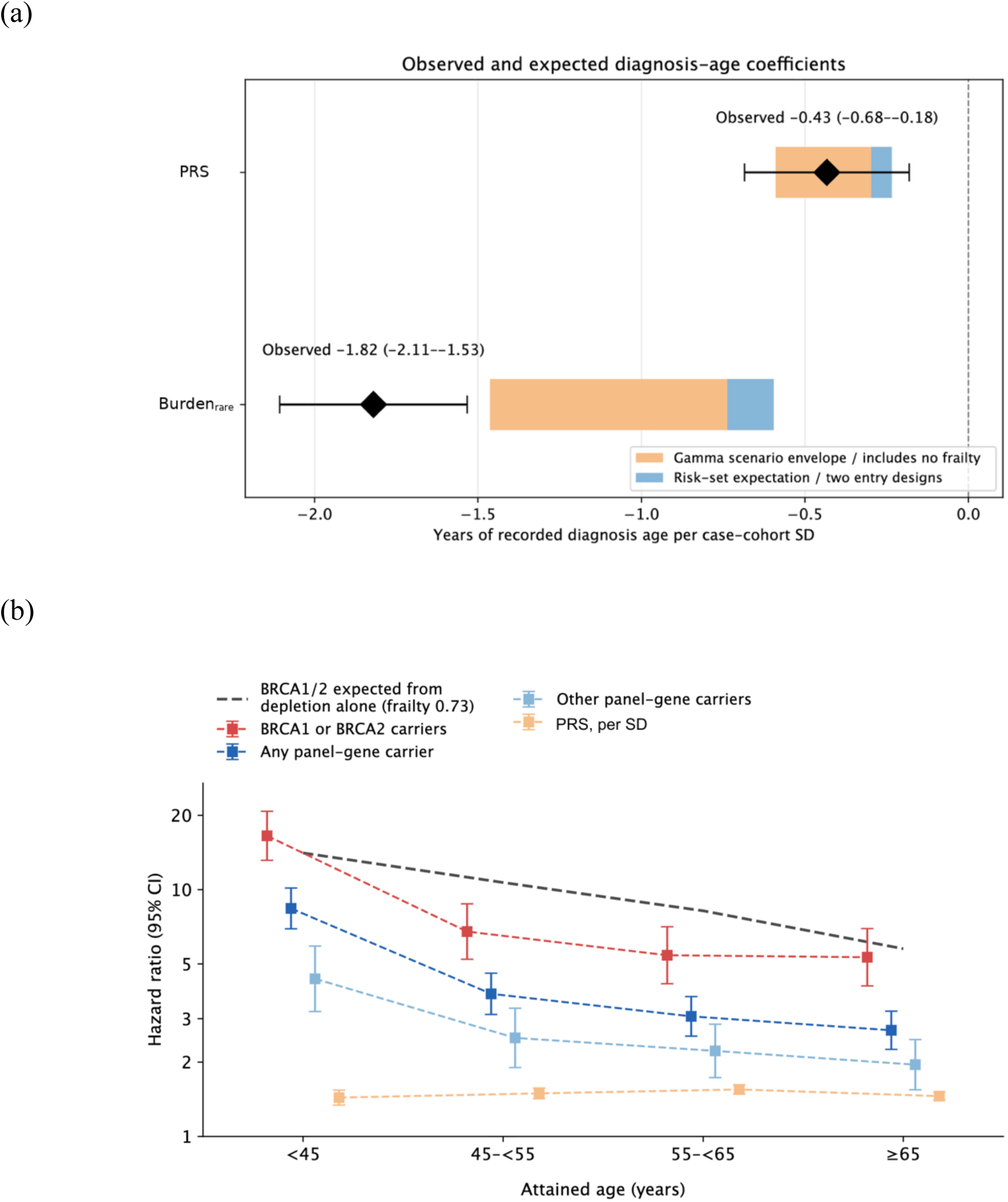
Diagnosis-age coefficients, age-specific hazard ratios, and frailty. (a) Diamonds show observed case-only diagnosis-age coefficients (Table 3). Blue bands show values expected from age-scale Cox risk sets under age-constant relative effects; orange bands span gamma-frailty simulations, including no frailty. (b) Points and 95% CIs show age-scale Cox HRs by attained age for PRS and carrier groups. The grey dashed line shows the BRCA1/BRCA2 decline predicted from high-risk-carrier depletion by the gamma-frailty model (variance 0.73).

## Discussion

In this All of Us cohort, PRS and Burden_rare_ each contributed to breast cancer risk prediction. Their joint association was sub-multiplicative on the odds-ratio scale. Among cases, greater Burden_rare_ was associated with younger recorded diagnosis age; the PRS association was smaller. Together, the scores explained only 2.63% of age-at-diagnosis variation.

The PRS odds ratio per standard deviation (1.55) was close to the 1.61 reported for the 313-variant score in prospective cohorts^[1]^, despite our use of EHR-defined case status. A weaker relative PRS association in BRCA1/BRCA2 carriers than in noncarriers was also reported in a carrier-stratified study^[6]^. In our exploratory gene-specific analysis, BRCA1 had the clearest negative interaction. Breast cancer subtype composition or the relevance of an overall PRS to different carrier groups could contribute to this difference^[5]^; subtype data were not analysed here. An interaction on the logistic scale does not establish biological antagonism or an interaction on the additive-risk scale.

The Burden_rare_ association with younger recorded diagnosis age is consistent with the earlier breast cancer onset reported for PALB2 and CHEK2 frameshift carriers in FinnGen^[8]^. The PRS association with age was consistent with the range expected from its breast cancer risk association alone.

The analysis was limited to genetically inferred European ancestry. Requiring a European posterior probability above 0.75 excluded some admixed participants, particularly near the European–Middle Eastern ancestry boundary; the results may not generalise to other ancestries. EHR-based phenotypes introduce uncertainty in diagnosis dates, leave some prevalent cases in the age analysis and may classify unrecorded cases as controls. Differences in EHR follow-up inflated the covariate-only AUC, so we interpret changes in AUC rather than its absolute value. The exploratory gene-specific interaction analyses were limited by sparse carrier counts, which required pooling several genes in the gene–PRS model. The external Burden_rare_ weights only approximate our qualifying variant definitions, the *STK11* weight is imprecise, and final manual adjudication of variants remains pending. Allele-fraction filtering cannot fully exclude clonal hematopoiesis. The original analysis plan was fixed after an earlier analysis in a related cohort; the full gene panel and gene–PRS diagnostics were developed later. Age-at-diagnosis findings are restricted to cases and may be affected by selection on case status^[21]^.

Common and rare germline variation both contributed to breast cancer risk prediction in this EHR-linked cohort. Burden_rare_ had a larger association with recorded diagnosis age among cases than PRS; neither score explained much of the variation in that age.

## Data availability

Individual-level data are available to registered researchers through the *All of Us* Researcher Workbench (controlled tier, CDR C2025Q4R6). Aggregate results that meet the dissemination policy and the analysis code are available at https://github.com/martina-yu/AoU_Comr.

## Ethics approval and consent to participate

The All of Us Research Program protocol and informed consent materials were approved by the All of Us Institutional Review Board, and participants provided informed consent for research use of their data. This study was a secondary analysis of data accessed through the controlled-tier Researcher Workbench. We did not contact participants or access directly identifiable information. Under the program’s data passport model, project-specific review by the All of Us Institutional Review Board was not required.

## Acknowledgments

We gratefully acknowledge All of Us participants for their contributions, without whom this research would not have been possible. We thank the National Institutes of Health’s All of Us Research Program for making the participant data examined in this study available.

